# Investigation of somatic mutations in human brains targeting genes associated with Parkinson’s disease

**DOI:** 10.1101/2020.05.15.20094722

**Authors:** Melissa Leija-Salazar, Alan Pittman, Katya Mokretar, Huw Morris, Anthony H. Schapira, Christos Proukakis

## Abstract

**Background:** Somatic mutations occur in neurons but their role in synucleinopathies is unknown.

**Aim:** We aimed to identify disease-relevant low-level somatic single nucleotide variants (SNVs) in brains from sporadic patients with synucleinopathies and a monozygotic twin carrying *LRRK2* G2019S, whose penetrance could be explained by somatic variation.

**Methods and Results:** We included different brain regions from 26 Parkinson’s disease (PD), 1 Incidental Lewy body, 3 multiple system atrophy cases and 12 controls. The whole SNCA locus and exons of other genes associated with PD and neurodegeneration were deeply sequenced using molecular barcodes to improve accuracy. We selected 21 variants at 0.33-5% allele frequencies for validation using accurate methods for somatic variant detection.

**Conclusions:** We could not detect disease-relevant somatic SNVs, however we cannot exclude their presence at earlier stages of degeneration. Our results support that coding somatic SNVs in neurodegeneration are rare, but other types of somatic variants may hold pathological consequences in synucleinopathies.

## Introduction

Synucleinopathies are disorders characterized by the pathological aggregation of α-synuclein^1^. Among synucleinopathies, Parkinson’s disease (PD) is the commonest disorder and is characterized predominantly by neurodegeneration of dopaminergic neurons in substantia nigra (SN)^2,3^. Somatic variation occurs in human brain and its role in neurodegeneration has started to be explored^4^. Current estimations of the occurrence of somatic variants in human brains suggest that single nucleotide variants (SNVs, or “point mutations”) could be the most prevalent form^5,6^. Somatic SNVs are reported to increase with age, where large genes or transcriptionally active genomic regions appear to be susceptible^7^. Somatic SNVs in coding regions of genes associated with synucleinopathies could contribute directly to these disorders, depending on the amount of affected cells and mechanisms of spread of the aetiological agent (see review^8^). The study of somatic SNVs has been facilitated by the latest technological improvements. Compared to single-cell studies, bulk-sequencing offers a cost-effective strategy to study somatic variation across tissues and brain regions of multiple individuals. The error rate of bulk-sequencing at low allele frequencies (AF) can be reduced by using molecular barcodes^9^. In this study, we used targeted sequencing in PD-associated genes from post-mortem human brains aimed for the detection of pathogenic somatic SNVs.

## Methods

Samples were obtained from the Parkinson’s UK and Queen Square brain banks. Patients gave informed consent and the study was approved by the local ethics committee. We evaluated 66 samples from multiple brain regions and 3 matched-blood samples, derived from 42 individuals with the following conditions: 26 PD, 12 control, 3 Multiple system atrophy (MSA) and 1 Incidental Lewy Body case. PD cases were sporadic, except for case 18, a manifesting *LRRK2* G2019S carrier, whose identical twin was non-penetrant^10^ (table S1) and somatic variation was suggested as an explanation for the discordance in the development of PD^11^. The mean onset age was 60.6±11 and mean disease duration 10.5±7 years.

We used a previously reported protocol for genomic DNA extraction^12^ and the Haloplex^HS^ method to prepare sequencing libraries. Details about the generation of artificial mosaics, the sequencing panel design, the customisation of library preparation and bioinformatic analysis are provided in table S2 and figure S1.

For amplicon sequencing, primers were designed with Primer3Plus^13^ to generate amplicons larger than 300 bp, targeting the variants of interest at >50 bp away from the primer annealing sites. Amplicons belonging to the same sample were pooled together before Nextera XT library preparation, following manufacturer instructions. Samples were pooled equimolarly before sequencing using a MiSeq v3 kit (600 cycles). The bioinformatic analysis is described in figure S2. Droplet digital PCR (ddPCR) assays were designed using Primer3plus, according to manufacturer. Bulk DNA from putamen, occipital, frontal cortex and cingulate gyrus was used for this analysis. The ddPCR conditions are described in table S3. Data analysis was performed in QuantaSoft Pro v1.0 following Bio-Rad guidelines.

## Results

### Validation of the methodology

“Artificial mosaics” were used to estimate the variant detection limit, sensitivity and false positive and negative rates. We were expecting 37 variants to be present within regions covered in artificial mosaics. We detected 95% of these variants at 1% AF and 87% at 0.5% AF (supplementary results).

We aimed to reduce to a minimum false positives at lower AF levels. We firstly counted ‘Potential false positives’ (PFP) in artificial mosaics at different AF thresholds. PFP comprised SNVs not recorded as expected mosaic variants, nor reported in dbSNP^14^. We observed 1.2x more PFP when the minimum AF was lowered from 1% to 0.5% (figure S3). Surecall showed greater sensitivity when compared to other variant callers (figure S3). To increase the specificity of our variant calling analysis, we filtered false positives visually, using fixed criteria to discard errors (figure S4). Surecall variants in mosaic 0.25% (at AF=0.25-5%) were analyzed on IGV. From the 114 variants analyzed, visual analysis could not discard 4 false positives. The highest AF was reported as 0.32%, therefore we set our detection limit at 0.33%. This filter allowed us to discard numerous false positives, but also increased the false negative rate. In the artificial mosaic sample carrying variants at 0.5%, Surecall detected 78% of the expected variants. After visual inspections, 46% of the expected variants remained, and false positives were completely discarded. The most common reason to filter real variants was their presence in only one paired-read orientation (strand-bias; figure S4B).

### Sample analysis

An explanation of our analysis is summarized in figure 1A. On the Haloplex^HS^ step, all samples were sequenced at an average 2541x. We focused on the detection of coding SNVs not reported before as common SNPs (population frequencies < 1%), to reduce the risk of calling low-level variants arising due to contamination. 31 variants in 23 samples passed the filtering step, but most of the variants detected (24 out of 31) had an average AF of 0.45%, close to the detection limit of our analysis. 21 variants in 18 samples were prioritized for validation, based on a ranking scale to select variants with a predictable role in disease (table S4). We generated amplicons to target the prioritized variants and sequenced those at even higher coverage (mean= 14,883x). To account for possible sequencing errors at the genomic positions of interest, we compared the amplicons from the interrogated sample with amplicons from controls (a commercial reference DNA and 6 samples showing a candidate variant in other parts of the genome). Two variants in samples 4SN and 34SN were validated, as these were detected at AFs close to the original analysis, and significantly different from the sequencing errors in controls (figure 1B). The variants were further confirmed by Mutect2 paired-analysis, using the reference DNA as a normal sample. However, these variants corresponded to rare heterozygous SNPs present in samples from our study. SN tissue was not available for further validation, but the AF at which the variants were detected was an indicative that the variants might be present in other brain regions when real^15^. ddPCR did not reveal the variants in the brain regions tested (figure 1C and D). In one of the assays, the presumably contaminated DNA was still available and the variant was confirmed only in this sample (figure 1C). To further examine cross-contamination, we recorded all mosaic variants from Surecall in 4SN, 34SN and control 1 (a sample used for demonstration purposes) at AF similar to the variants of interest. The mosaic variants were compared to germline variants from samples where the contamination was suspected to come from (in the case of control 1, a non-related sample or control 2). While control 1 showed fewer mosaic variants, not matched with control 2 germline variants, the presumably contaminated samples showed numerous mosaic variants matched with germline variants from samples where the contamination came from (p<0.0001, linear regression; figure S5).

**Figure 1.**
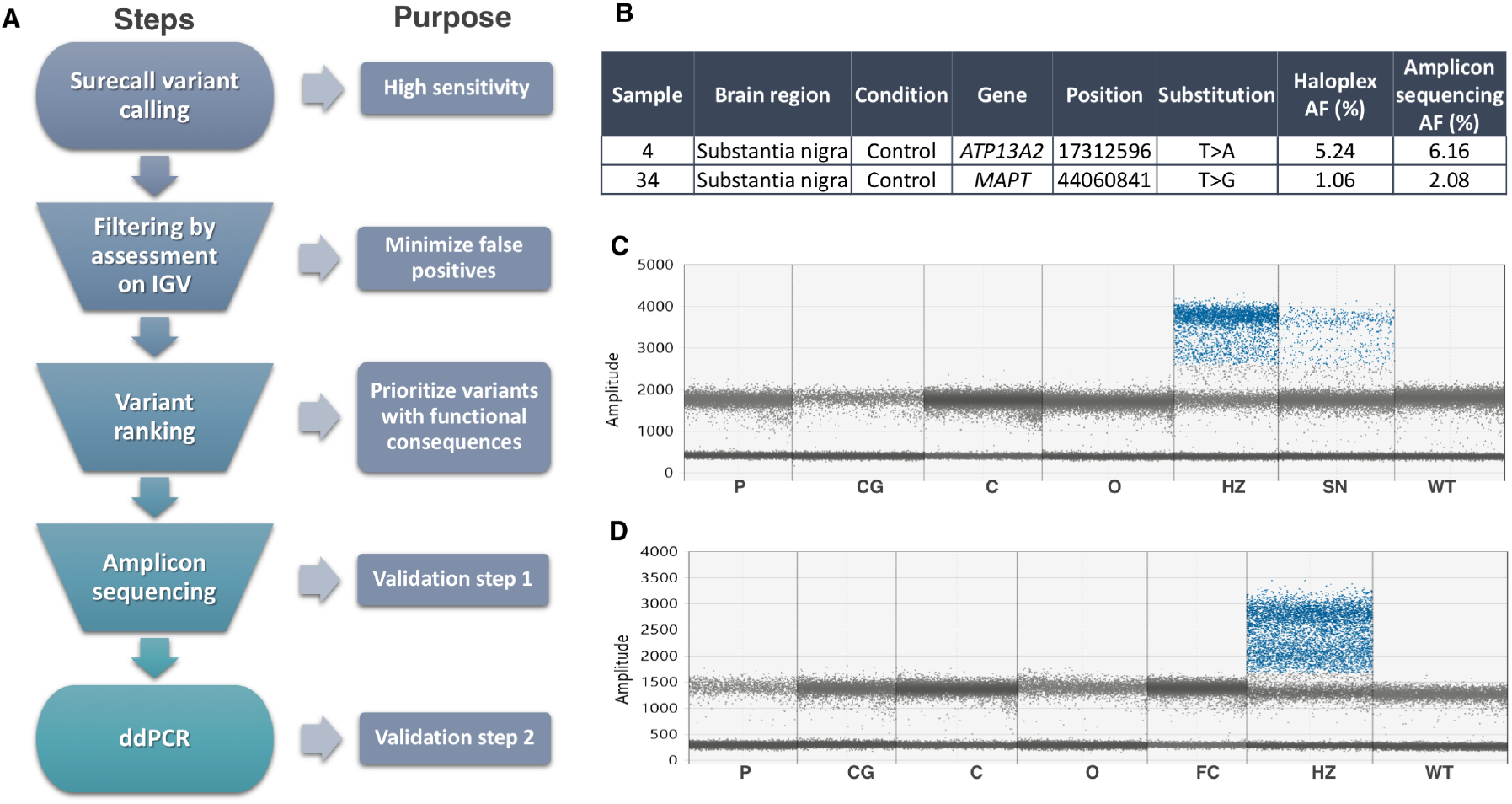
Summary of methods and results. (A) Somatic variant calling workflow explained step by step. (B) Validated variants by Amplicon sequencing (AF=allele frequency). (C) ddPCR assay for the *ATP13A2* variant did not reveal its presence in additional brain regions of sample 4, nor in control DNA (WT). Sample 32SN (presumable contaminant) showed the variant at heterozygous levels (HZ). The presumably contaminated sample 4SN used in Haloplex^HS^ and Amplicon sequencing assays showed a mutant signal at AF ~6%. (D) ddPCR assay for the *MAPT* variant did not reveal its presence in additional brain regions of sample 34, nor in control DNA (WT). Sample 22SN (presumable contaminant) showed the variant at heterozygous levels (HZ). Codes for brain regions tested: SN=substantia nigra, P=putamen, CG=cingulate gyrus, C=cerebellum, O=occipital.

## Discussion

Previous work from our group could not detect somatic SNVs in *SNCA* exons at AF above 5% in cerebellum, frontal cortex and SN of sporadic PD patients^16^. In this study, we expanded our search to other PD-genes. We excluded as many cases as possible with long disease duration and late-onset, as somatic variants playing a role in disease are hypothesized to be less likely to occur in these cases^16,17^. We included a patient carrying a *LRRK2* G2019S mutation, who had a phenotypically discordant monozygotic twin and where somatic variation could have played a role in penetrance. We used a highly sensitive approach to detect low-level variants in the genes of interest, by firstly combining deep sequencing coverage and molecular barcodes, followed by amplicon sequencing at higher coverage and ddPCR as validation steps^18^. We could not detect somatic SNVs in PD-associated genes at AF higher than 0.33%. Similar to our results, a recent report could not identify somatic SNVs at AF above 0.5% in familial PD-genes from brains with Lewy body disorders (n=20), using similar methodologies and higher sequencing coverage^19^. Previous studies using Haloplex^HS^ reported variant detection at AF above 0.2%, further supporting that our analysis was close to the detection limits of this methodology^19-21^. We focused on refining the analysis to mainly discard false positives. Our filtering criteria were tailored to discard sequencing artefacts, similar to other studies using Haloplex and common sequencing datasets^22-26^. Advantages of visual analysis are the comprehensive analysis for each variant, easy implementation across datasets; however, it can become labour-intensive. Our results demonstrate the difficulties of SNV detection at low AF, due to low-level contamination and false positives, even when using molecular barcodes.

Challenges of somatic variant studies are not only technical, but also related to the stochastic nature of the variants. According to a previous hypothesis where neurons carrying somatic variation may be the most vulnerable and first to degenerate, we selected for patients with disease duration as short as possible (~10 years)^16^. When studying neurodegeneration in post-mortem brains, only the latest stages of the disease are being portrayed and, perhaps, events involved in disease development are missed.

Our data combined with work discussed above, suggest that coding somatic SNVs in PD-associated genes are uncommon. In Alzheimer’s disease, two brain somatic SNVs were found in 72 sporadic AD-patients^27^. When using molecular barcodes, two brain somatic SNVs were found in AD-associated genes of 98 patients ^28^, whereas no somatic SNVs in familial AD-genes were found in 20 patients^19^. Somatic SNVs in *APP* were reported in AD in the context of the novel mechanism of recombination leading to “genomic cDNA”^29^. Recently, 14 out of 52 AD-patients analyzed by deep exome sequencing harboured exonic somatic mutations in genes involved in tau phosphorylation, but not familial AD genes^30^. This contrasts with somatic CNVs, with *SNCA* gains in PD nigral dopaminergic and cortical neurons^31, 32^.

In summary, our study could not detect coding somatic SNVs at AF above 0.33% when analysing PD-associated genes from brain samples. Reaching lower AF to detect late somatic variant events using bulk-tissue requires an even larger sequencing effort, and it is complicated by the common presence of contamination and sequencing errors. Sequencing of dopaminergic single-nuclei should give enough resolution to describe somatic variants in cells mainly affected by PD (dopaminergic neurons). Additional studies can be aimed to explore other types of somatic variations or other mechanisms by which somatic SNVs outside PD-associated genes could play detrimental roles in neurodegeneration.

## Data Availability

All data will be freely available on June 1st.

## Data availability statement

The data that support the findings will be openly available after July 31st, 2020 in the European Nucleotide Archive (ENA by EMBL-EBI) with the primary accession number of PRJEB36518.

## Authors Roles

MLS conducted the experiments, analyzed the data, wrote, revised and submitted this manuscript. AP participated in the experimental design and data analysis. KM participated in the experimental design and performed initial experiments. HM and AS participated in the design of the study. CP conceived and designed the study, contributed to interpret the data and revised the final version of the manuscript. All authors read and approved the final version of the manuscript.

## Acknowledgments

We are grateful to all individuals who donated their brains for research. CP received funding from the Michael J. Fox Foundation for Parkinson’s disease research and MS was partly funded by CONACYT. AS is funded by the UK Medical Research Council, and the Kattan Trust. Queen Square Brain Bank is supported by the Reta Lila Weston Institute for Neurological Studies and the Medical Research Council UK. The Parkinson’s UK Tissue Bank is funded by Parkinson’s UK, a charity registered in England and Wales (258197) and in Scotland (SC037554).

